# QT Interval Prolongation and Torsade De Pointes in Patients with COVID-19 treated with Hydroxychloroquine/Azithromycin

**DOI:** 10.1101/2020.04.27.20074583

**Authors:** Ehud Chorin, Lalit Wadhwani, Silvia Magnani, Matthew Dai, Eric Shulman, Charles Nadeau-Routhier, Robert Knotts, Roi Bar-Cohen, Edward Kogan, Chirag Barbhaiya, Anthony Aizer, Douglas Holmes, Scott Bernstein, Michael Spinelli, David Park, Carugo Stefano, Larry Chinitz, Lior Jankelson

## Abstract

**Background:** The emergence of the COVID-19 pandemic has resulted in over two million affected and over 150 thousand deaths to date. There is no known effective therapy for the disease. Initial reports suggesting the potential benefit of Hydroxychloroquine/Azithromycin (HY/AZ) have resulted in massive adoption of this combination worldwide. However, while the true efficacy of this regimen is unknown, initial reports have raised concerns regarding the potential risk of QT prolongation and induction of torsade de pointes (TdP).

**Methods:** This is a multicenter retrospective study of 251 patients with COVID-19 treated with HY/AZ. We reviewed ECG tracings from baseline and until 3 days after completion of therapy to determine the progression of QTc and incidence of arrhythmia and mortality.

**Results:** QTc prolonged in parallel with increasing drug exposure and incompletely shortened after its completion. Extreme new QTc prolongation to > 500 ms, a known marker of high risk for TdP had developed in 15.9% of patients. One patient developed TdP requiring emergent cardioversion. Seven patients required premature termination of therapy. The baseline QTc of patients exhibiting QTc prolongation of > 60 ms was normal.

**Conclusion:** The combination of HY/AZ significantly prolongs the QTc in patients with COVID-19. This prolongation may be responsible for life threating arrhythmia in the form of TdP. This risk mandates careful consideration of HY/AZ therapy in lights of its unproven efficacy. Strict QTc monitoring should be performed if the regimen is given.

## Introduction

Clusters of severe respiratory illness caused by SARS-CoV-2 virus emerged in Wuhan, China, in late 2019 [1]. As of April 16 2020, a total of 2,193,666 individuals in 208 countries were reported to be infected by SARS-CoV-2, which causes coronavirus disease 2019 (COVID-19), with more than 158,000 deaths [2]. The evidence supporting effective drug therapy for COVID-19 is limited. In vitro studies have suggested that Hydroxychloroquine alone and in combination with Azithromycin could be a viable treatment for COVID-19 [3, 4]. A small, controversial study in France enrolling 26 treated patients and 16 non-randomized controls showed that Hydroxychloroquine in combination with Azithromycin (HY/AZ) shortened the time to resolution of viral shedding of SARS-CoV-2 [5]. Based on this, clinicians in many countries have begun using these medications in clinical practice, and multiple randomized trials are ongoing [6]. However, HY and AZ have each been independently shown to increase the risk for QT interval prolongation, drug-induced torsade de pointes (TdP), and drug induced sudden cardiac death (SCD) [7–10]. We recently reported a frequent and significant prolongation of the QT interval in a preliminary series of 84 adult patients with SARS-CoV-2 infection treated with HY/AZ combination [11]. Here we report a multicenter study evaluating the effects of HY/AZ on the QT interval and the arrhythmic risk in patients with SARS-CoV-2 infection.

## Methods

This is a multicenter, retrospective study performed at NYU Langone Medical Center, New York, USA and San Paolo Hospital, Milan, Italy. We included 251 consecutive adult patients hospitalized at NYU Langone Medical Center (211 patients) and at San Paolo Hospital (40 patients) with COVID-19 disease and treated with the combination of HY/AZ. 84 of the 211 patients from the NYU site were reported in our recent publication [11]. Medical records were reviewed to obtain baseline characteristics, laboratory data, and serial ECGs. Hydroxychloroquine was given orally at 400 mg BID for one day (loading dose) followed by 200 mg BID for 4 days. Azithromycin was given orally at a dose of 500 mg daily for 5 days. For QTc measurement, five cardiologists trained and experienced in QT measurement performed all electrocardiographic measurements. For quality assurance, QT and RR measurements were validated by a senior electrophysiologist experienced in QT measurements. QTc was calculated from the QT and RR intervals using the Bazetts’ formula.

The closing date of follow-up was April 15^th^ 2020. Collected data on the closing date included arrhythmic events and mortality. The primary endpoints included absolute QTc >500ms, a known marker of high risk for malignant arrhythmia and sudden cardiac death [12–14], and QTc prolongation by >60 ms, another high risk marker [15].

## Statistical analysis

Statistical analysis was performed using IBM SPSS Statistics 26, and figures were constructed using GraphPad Prism 8. Continuous variables are expressed as mean ± standard deviation, and categorical variables are expressed as percentages. Normality of data samples was assessed using Shapiro-Wilk test. Two sample hypothesis testing for continuous variables was performed using Student’s *t*-test if samples had normal distributions, Mann-Whitney *U* test if samples did not have normal distributions, or paired samples *t*-test for paired samples. Two sample hypothesis testing for categorical variables was performed using Fisher’s exact test. For Figure 2, one sample t-test (if samples were normally distributed) or one sample Wilcoxon signed rank test (if samples were not normally distributed) were performed to compare each sample against a delta QTc of 0 ms (i.e. no change from baseline), and p values were adjusted using the Holm-Bonferroni method to α < 0.05. For Tables 2 and 3, univariate and multivariate logistic regression was performed to identify predictors of maximum QTc >500 ms and ΔQTc > 60 ms.

The study was performed according to the NYU Institutional Review Board and Quality Improvement initiative and the University Hospital of Milan Institutional Review Board guidance in accordance with the ethical standards laid down in the 1964 Declaration of Helsinki and its later amendments.

## Results

We included 251 patients in our cohort with a maximal follow up of 8 days and mean ECG follow up time of 5.6 ± 2.2 days. The clinical and epidemiological characteristics are presented in Table 1. The median age was 64±13 years and 75% were male. 44 (17.5%) patients died of respiratory or multi-organ failure. One patient with extreme QTc prolongation developed TdP and required emergent cardioversion (Figure 1), representing an arrhythmic risk of 0.4%. Overall, QTc interval prolonged from a baselines of 439 ± 29 ms to a maximal value of 474 ± 37 ms (p<0.001) which occurred on day 4.2 ± 2.1 of therapy. The daily QTc distribution and change are presented in Figure 2. Of note, in 40 (15.9%) patients extreme QTc prolongation was observed. In this high risk group, QTc increased from a baseline of 457.6 ± 33.2 to 535.0 ± 30.2 ms (p<0.001). Of those, in 7 patients extreme QTc prolongation triggered discontinuation of therapy prematurely on day 3. QTc prolongation (ΔQTc) of > 60 ms, occurred in 51 (20.3%) patients, from a baseline of 426.3 ± 29.6 to 513.3 ± 40.2 ms. On multivariate analysis, baseline QTc, creatinine during maximal QTc and co-administration of Amiodarone were significant predictors of extreme QTc prolongation (Table 2). Patients with extreme QTc prolongation had greater frequency of kidney disease, congestive heart failure, and Amiodarone exposure (Supplementary table 1). The predictors of ΔQTc > 60 ms were shorter baseline QTC, baseline creatinine and co-administration of Amiodarone (Table 3). Patients exhibiting ΔQTc > 60 ms had a lower body weight, greater frequency of coronary artery and kidney disease and more Amiodarone exposure (Supplementary table 2).

**Table 1.** Baseline patient characteristics

|  |  |
| --- | --- |
| Age (years) | 64 ± 13 |
| Gender (% male) | 75% |
| Weight (kg) | 86.0 ± 17.9 |
| Coronary artery disease | 12% |
| Hypertension | 54% |
| Chronic kidney disease | 11% |
| Diabetes mellitus | 27% |
| Chronic obstructive pulmonary disease | 7% |
| Congestive heart failure | 3% |
| Creatinine at initiation (mg/dL) | 1.2 ± 0.9 |
| Creatinine at max QTc (mg/dL) | 1.6 ± 1.5 |
| CrCl at initiation (mL/min) | 84 ± 43 |
| CrCl at max QTc (mL/min) | 80 ± 52 |
| Abnormal liver function tests at initiation | 21% |
| Abnormal liver function tests at max QTc | 38% |
| Potassium at baseline (mEq/L) | 4.1 ± 0.6 |
| Potassium at max QTc (mEq/L) | 4.2 ± 0.5 |
| QTc-prolonging medications |  |
| Psychiatric medications | 12% |
| Anti-microbials | 10% |
| Amiodarone | 9% |
| Baseline QTc (ms) | 439 ± 29 |
| Maximum QTc (ms) | 474 ± 37 |
| Maximum delta QTc (ms) | 35 ± 36 |
| Day of maximum QTc | 4.2 ± 2.1 |

**Table 2.** Predictors of Maximum QTc >=500ms

| Univariate Logistic Regressions |  |  |  |
| --- | --- | --- | --- |
| Variable | *p* value | OR | 95% CI |
| Age (yrs) | 0.47 | 1.01 | .98 - 1.04 |
| Gender (% male) | 0.35 | 1.48 | .65 - 3.4 |
| Weight (kg) | 0.14 | 0.99 | .97 - 1.01 |
| Coronary artery disease | 0.26 | 1.70 | .68 - 4.28 |
| Hypertension | 0.89 | 1.05 | .54 - 2.04 |
| Chronic kidney disease | 0.06 | 2.35 | .95 - 5.81 |
| Diabetes mellitus | 0.29 | 1.47 | .72 - 3. |
| Chronic obstructive pulmonary disease | 0.99 | 1.00 | .28 - 3.6 |
| Congestive heart failure | 0.01 | 7.23 | 1.56 - 33.6 |
| Creatinine at initiation (mg/dL) | 0.01 | 1.78 | 1.19 - 2.66 |
| Creatinine at max QTc (mg/dL) | <0.01 | 1.33 | 1.1 - 1.61 |
| CrCl at initiation (mL/min) | 0.03 | 0.99 | .98 - 1. |
| CrCl at max QTc (mL/min) | <0.01 | 0.99 | .98 - 1. |
| Abnormal liver function tests at initiation | 0.87 | 1.07 | .48 - 2.43 |
| Abnormal liver function tests at max QTc | 0.48 | 0.78 | .39 - 1.55 |
| Potassium at baseline (mEq/L) | 0.62 | 0.85 | .43 - 1.65 |
| Potassium at max QTc (mEq/L) | 0.70 | 0.86 | .41 - 1.83 |
| Psychiatric medications | 0.26 | 1.70 | .68 - 4.28 |
| Anti-microbials | 0.85 | 0.90 | .29 - 2.75 |
| Amiodarone | <0.01 | 4.71 | 1.91 - 11.65 |
| Baseline QTc (ms) | <0.01 | 1.03 | 1.02 - 1.04 |
| Multivariate Logistic Regression |  |  |  |
| Variable | p value | OR | 95% CI |
| Congestive heart failure | 0.62 | 1.58 | .27 - 9.37 |
| Creatinine at max QTc (mg/dL) | 0.03 | 1.26 | 1.03 - 1.55 |
| Amiodarone | 0.04 | 2.99 | 1.08 - 8.31 |
| Baseline QTc (ms) | <0.01 | 1.03 | 1.01 - 1.04 |

**Table 3.** Predictors of Delta QTc >=60ms

| Univariate Logistic Regressions |  |  |  |
| --- | --- | --- | --- |
| Variable | p value | OR | 95% CI |
| Age (yrs) | 0.13 | 1.02 | 1. - 1.04 |
| Gender (% male) | 0.70 | 1.15 | .56 - 2.37 |
| Weight (kg) | 0.01 | 0.98 | .96 - 1. |
| Coronary artery disease | 0.07 | 2.19 | .95 - 5.05 |
| Hypertension | 0.17 | 1.55 | .83 - 2.89 |
| Chronic kidney disease | 0.11 | 2.05 | .86 - 4.86 |
| Diabetes mellitus | 0.32 | 1.40 | .72 - 2.7 |
| Chronic obstructive pulmonary disease | 0.63 | 0.73 | .2 - 2.63 |
| Congestive heart failure | 0.63 | 1.51 | .29 - 8.03 |
| Creatinine at initiation (mg/dL) | 0.04 | 1.38 | 1.02 - 1.86 |
| Creatinine at max QTc (mg/dL) | 0.02 | 1.25 | 1.05 - 1.5 |
| CrCl at initiation (mL/min) | 0.04 | 0.99 | .98 - 1. |
| CrCl at max QTc (mL/min) | <0.01 | 0.99 | .98 - 1. |
| Abnormal liver function tests at initiation | 0.20 | 0.59 | .26 - 1.34 |
| Abnormal liver function tests at max QTc | 0.43 | 0.79 | .43 - 1.43 |
| Potassium at baseline (mEq/L) | 0.22 | 1.49 | .79 - 2.79 |
| Potassium at max QTc (mEq/L) | 0.34 | 1.40 | .7 - 2.8 |
| Psychiatric medications | 0.67 | 1.22 | .49 - 3.03 |
| Anti-microbials | 0.80 | 1.14 | .43 - 2.99 |
| Amiodarone | 0.01 | 3.31 | 1.36 - 8.05 |
| Baseline QTc (ms) | <0.01 | 0.98 | .97 - .99 |
| Multivariate Logistic Regression |  |  |  |
| Variable | *p* value | OR | 95% CI |
| Weight (kg) | 0.11 | 0.98 | .96 - 1. |
| Creatinine at initiation (mg/dL) | 0.02 | 1.73 | 1.1 - 2.7 |
| Amiodarone | <0.01 | 5.47 | 1.87 - 16.02 |
| Baseline QTc (ms) | <0.01 | 0.97 | .96 - .99 |

**Figure 1:**
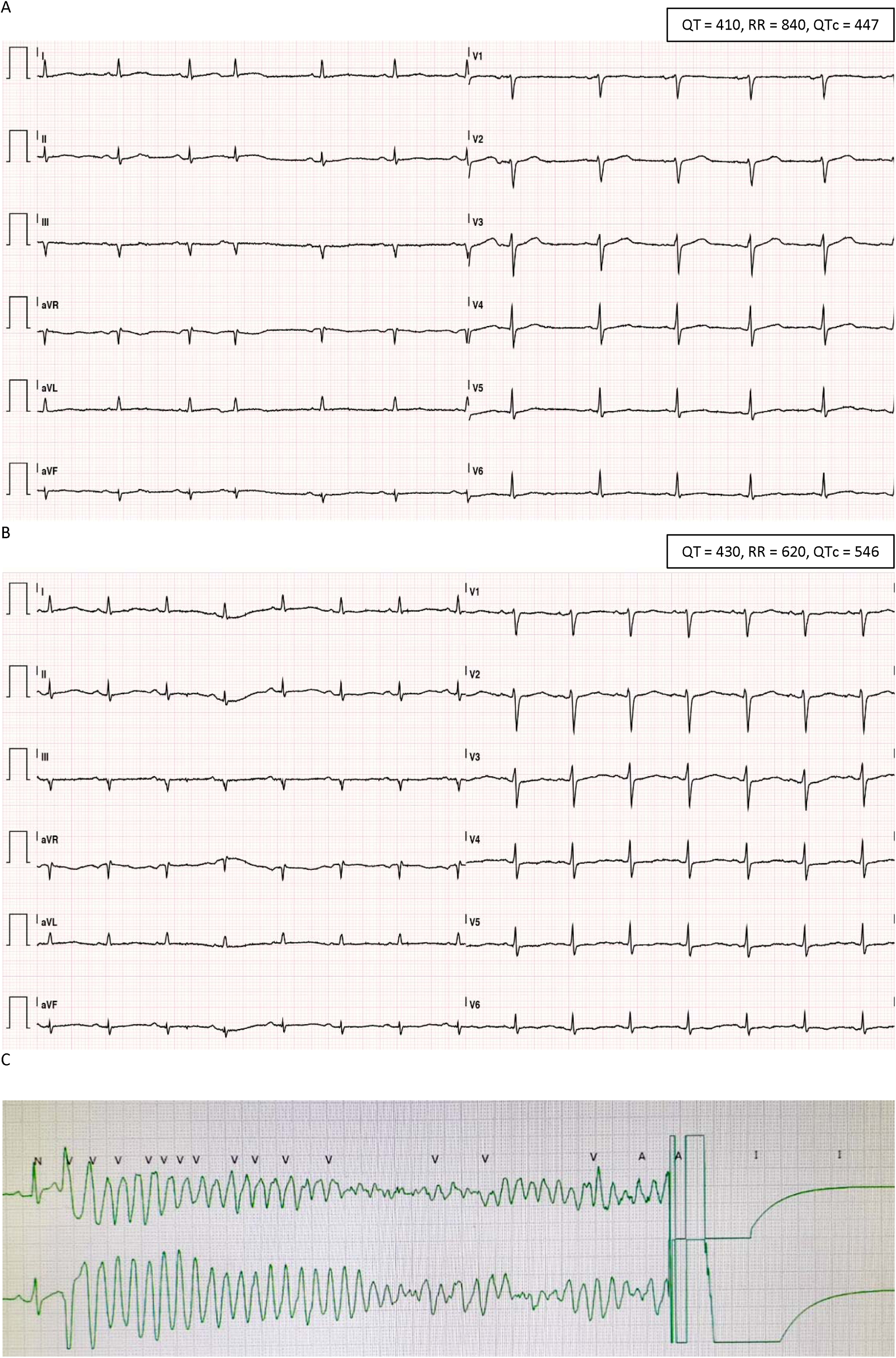
QTc prolongation and Torsdaes de point. A. baseline ECG before the initiation of HY/AZ. QTc = 447 ms. B. ECG at day 4 of HY/AZ, QTc prolonged to 582 ms. C. The same night, HY/AZ was stopped but the patient developed TdP requiring cardioversion.

**Figure 2:**
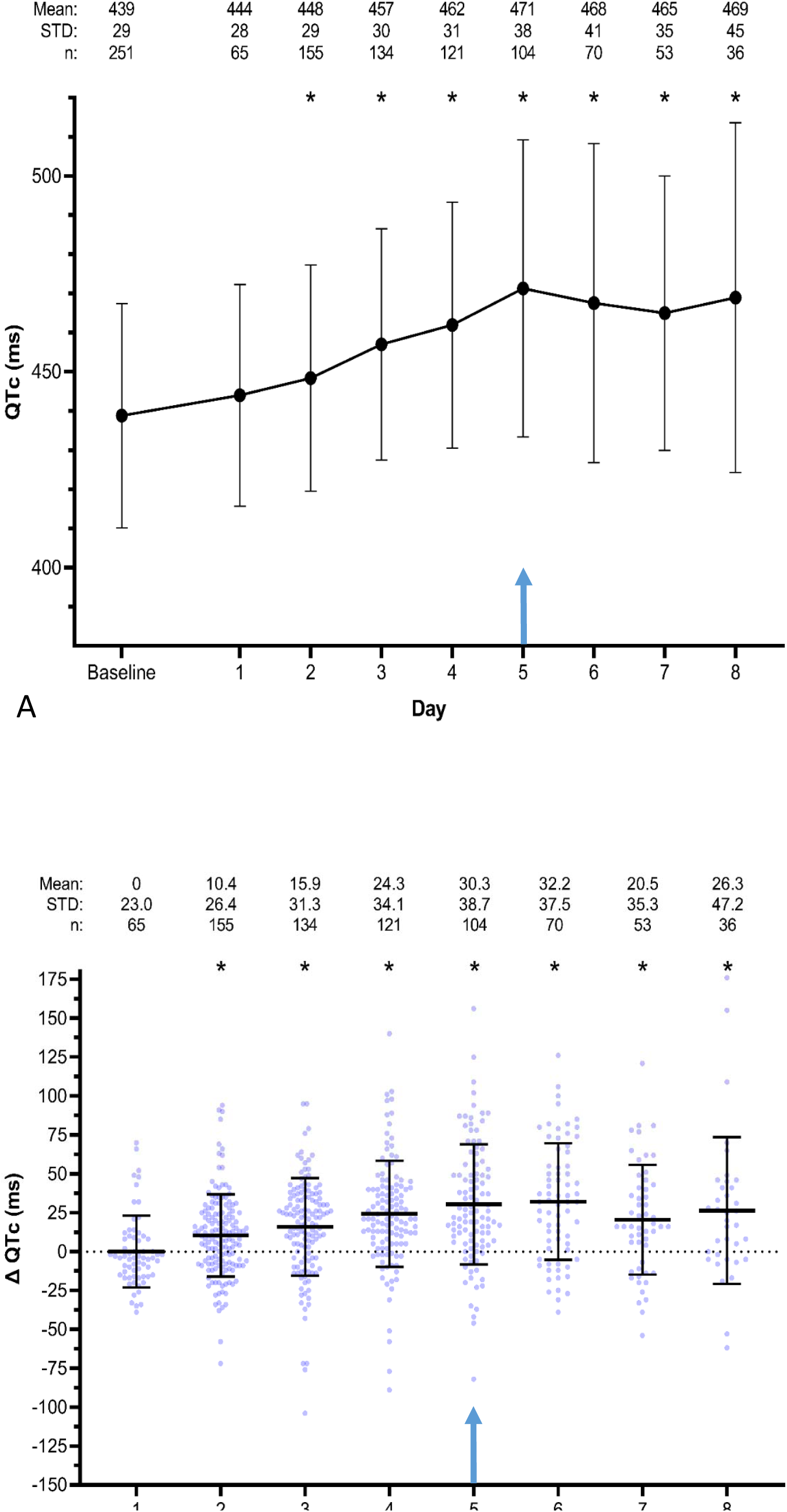
A. Daily absolute QTc in patients treated with HY/AZ and B. change in QTc by day. Number of patients, mean QTc +/- SD are presented at each day. * represents p<0.01 for the comparison with baseline QTc. Blue arrows indicate end of HY/AZ therapy.

**Figure 3:**
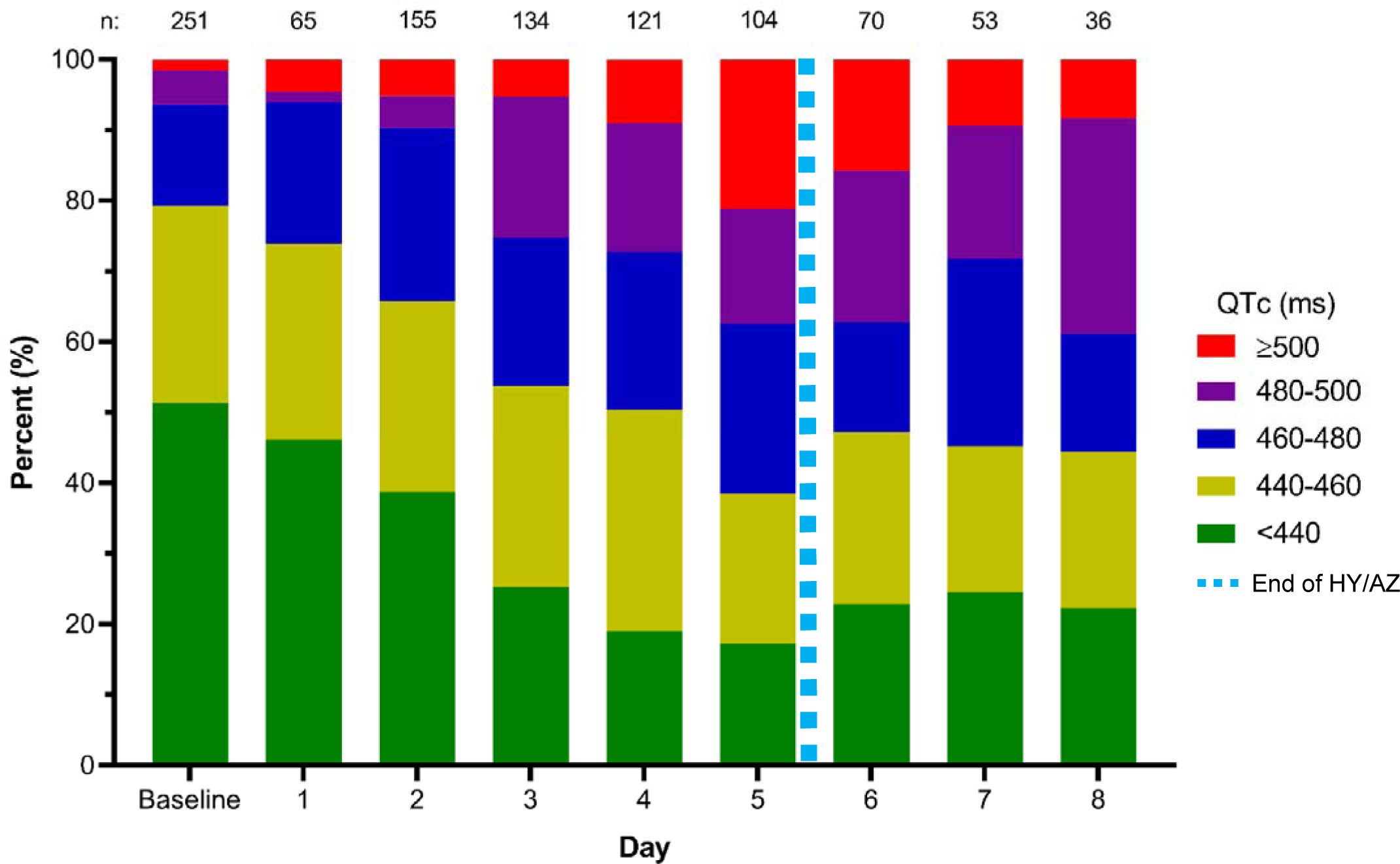
A. Distribution of QTc ranges by day of therapy. Note that therapy was given on days 1–5 (dashed line).

## Discussion

Drug-induced QT prolongation is an important substrate for TdP, a potentially lethal polymorphic ventricular tachycardia [16]. In our multicenter study of 251 patients with COVID-19, we found high incidence of QTc prolongation, resulting in at least one documented TdP at a rate of 0.4%. To put this incidence of TdP in perspective, the risk of TdP induction by Sotalol, a non-selective beta blocker used for the treatment of arrhythmia is estimated at 0.1% [17]. Due to this risk, Sotalol is introduced under ECG monitoring in the hospital for at least 3 days. In our study, we observed QTc prolongation in parallel with increasing HY/AZ exposure, which partially shortened after medication cessation. Baseline QTc, creatinine level and co-administration of Amiodarone were significant predictors of extreme QTc prolongation and ΔQTc > 60 ms. Previous information on the potential proarrhythmic effect of the combination Chloroquine/Hydroxychloroquine and Azithromycin is limited. In a randomized, placebo-controlled parallel trial in 116 young healthy controls receiving Chloroquine alone or in combination with Azithromycin, co-administration increased the QTc interval (Fridericia) by up to 14 ms [18]. However, the risk for drug induced TdP is substantially higher in hospitalized patients. This, is due to greater prevalence of other risk factors for TdP, including older age, presence of underlying heart disease, electrolyte disturbances and co-treatment with other QT prolonging medications [19–21]. Indeed, in a recent study assessing Chloroquine therapy in COVID-19 patients, extreme QT prolongation and excess cardiac mortality in the higher dose arm led to premature interruption of the regimen [22]. Concordantly, we recently published our preliminary experience on the dynamics of the QTc interval in patients with COVID-19. We found that 11% of patients developed extreme QTc prolongation greater than 500 ms. [11]. In the current study, we found an even higher proportion of patients with extreme QTC prolongation, 15.9%, with at least one TdP event requiring cardioversion. Another 7 patients who developed extreme QTc prolongation by day 3 had the treatment stopped, possibly preventing additional arrhythmic events. The alarming proportion of COVID-19 patients developing extreme QT prolongation with HY/AZ therapy in our study can be explained by the specific characteristics of this population, which includes older age, greater prevalence of underlying and acute heart disease, renal failure and co-administration of additional QT prolonging medications, particularly Amiodarone.

Recently published guidance statements addressing QTc surveillance and arrhythmic risk in COVID-19 patients are based on LQTS risk stratification principals and include pretreatment assessment of the QTc, considering stopping other QTc prolonging medications, and providing special attention to those with highest risk features [23–25]. Yet, our findings suggest that risk stratification of patients with COVID19 may be more complex. For example, we found that the baseline QTc in patients with ΔQTc > 60 ms was only 426.3 ± 29.6 ms, deep within the “normal” QTc range. Additionally, it is important to consider that even careful monitoring of the QT interval may only partially mitigate the risk for TdP. This is because arrhythmia often occurs in the setting of sudden, intermittent changes in the R-R interval, such as when PVCs, APCs or pauses occur. In these cases, TdP can present even if the QTc in only mildly prolonged at baseline [26]. We therefore suggest that individual risk/benefit assessment should be applied before treating with HY/AZ. We recommend daily ECG monitoring, with reassessment of the therapy if high risk markers appear (QTc >500 ms or ΔQTc > 60 ms). Finally, we observed only partial resolution of the QTc at 3 days after completion of therapy. This may be attributed to the prolonged half-life of Hydroxychloroquine, which is approximately 20 days. This finding requires special attention when considering discharging patients receiving HY/AZ or if outpatient treatment with HY/AZ is planned.

## Conclusions

Treatment of COVID-19 with HY/AZ prolongs the QTc to an extreme degree in a significant proportion of patients, increasing the risk for TdP. Risk/benefit considerations should be carefully and individually evaluated and preventive measures should be applied when using this regimen.

## Limitations

This is a retrospective study. Relatively short follow-up time after HY/AZ regimen completion was available. We did not assess serum drug concentrations.

## Data Availability

data will be made available as decided by the DSC of the study

---

**Supplementary Table 1.** Characteristics of patients with maximum QTc ≥500ms

|  | QTc <500ms<br>(n = 211) | QTc ≥500ms<br>(n = 40) | *p* |
| --- | --- | --- | --- |
| Age (yrs) | 64 ± 13 | 65 ± 15 | 0.58 |
| Gender (% male) | 74% | 83% | 0.32 |
| Weight (kg) | 86.6 ± 17.8 | 82.9 ± 18.4 | 0.27 |
| Coronary artery disease | 10% | 18% | 0.28 |
| Hypertension | 54% | 55% | 1 |
| Chronic kidney disease | 9% | 20% | 0.05 |
| Diabetes mellitus | 25% | 35% | 0.24 |
| Chronic obstructive pulmonary disease | 7% | 8% | 1 |
| Congestive heart failure | 1% | 10% | 0.01 |
| Creatinine at initiation (mg/dL) | 1.1 ± 0.5 | 1.8 ± 2.0 | 0.02 |
| Creatinine at max QTc (mg/dL) | 1.4 ± 1.3 | 2.3 ± 1.9 | <0.01 |
| CrCl at initiation (mL/min) | 87 ± 43 | 72 ± 41 | 0.04 |
| CrCl at max QTc (mL/min) | 84 ± 52 | 56 ± 43 | <0.01 |
| Abnormal liver function tests at initiation | 21% | 24% | 0.67 |
| Abnormal liver function tests at max QTc | 39% | 28% | 0.63 |
| Potassium at baseline (mEq/L) | 4.2 ± 0.6 | 4.1 ± 0.6 | 0.38 |
| Potassium at max QTc (mEq/L) | 4.2 ± 0.5 | 4.2 ± 0.6 | 0.47 |
| Psychiatric medications | 10% | 18% | 0.28 |
| Anti-microbials | 10% | 10% | 1 |
| Amiodarone | 6% | 25% | <0.01 |
| Baseline QTc (ms) | 435 ± 26 | 457 ± 34 | <0.01 |
| Maximum QTc (ms) | 461 ± 23 | 534 ± 30 | <0.01 |
| Maximum delta QTc (ms) | 26 ± 28 | 77 ± 40 | <0.01 |
| Day of maximum QTc | 4.0 ± 2.0 | 4.6 ± 1.8 | 0.05 |

**Supplementary Table 2.** Characteristics of patients with Delta QTc ≥60ms

|  | Delta QTc <60ms<br>(n = 200) | Delta QTc ≥60ms<br>(n = 51) | *p* |
| --- | --- | --- | --- |
| Age (yrs) | 63 ± 13 | 66 ± 13 | 0.18 |
| Gender (% male) | 75% | 78% | 0.72 |
| Weight (kg) | 87.3 ± 18.6 | 81.0 ± 14.0 | 0.03 |
| Coronary artery disease | 10% | 20% | 0.05 |
| Hypertension | 52% | 63% | 0.16 |
| Chronic kidney disease | 9% | 18% | 0.08 |
| Diabetes mellitus | 25% | 33% | 0.29 |
| Chronic obstructive pulmonary disease | 8% | 6% | 1 |
| Congestive heart failure | 3% | 4% | 0.63 |
| Creatinine at initiation (mg/dL) | 1.1 ± 0.7 | 1.5 ± 1.6 | 0.12 |
| Creatinine at max QTc (mg/dL) | 1.4 ± 1.3 | 2.1 ± 1.8 | <0.01 |
| CrCl at initiation (mL/min) | 87 ± 44 | 74 ± 38 | 0.06 |
| CrCl at max QTc (mL/min) | 85 ± 53 | 60 ± 40 | <0.01 |
| Abnormal liver function tests at initiation | 23% | 16% | 0.34 |
| Abnormal liver function tests at max QTc | 40% | 29% | 0.6 |
| Potassium at baseline (mEq/L) | 4.1 ± 0.6 | 4.3 ± 0.7 | 0.54 |
| Potassium at max QTc (mEq/L) | 4.2 ± 0.5 | 4.3 ± 0.6 | 0.63 |
| Psychiatric medications | 11% | 14% | 0.62 |
| Anti-microbials | 10% | 12% | 0.8 |
| Amiodarone | 7% | 20% | 0.01 |
| Baseline QTc (ms) | 442 ± 27 | 425 ± 29 | <0.01 |
| Maximum QTc (ms) | 463 ± 27 | 512 ± 40 | <0.01 |
| Maximum delta QTc (ms) | 21 ± 23 | 87 ± 26 | <0.01 |
| Day of maximum QTc | 4.0 ± 2.0 | 4.7 ± 1.8 | 0.01 |

